# Immediate and mid-term effect of a natural topical product in patients with musculoskeletal pain: randomized, double-blinded, and placebo-controlled clinical trial

**DOI:** 10.1101/2021.11.04.21265860

**Authors:** Silvia Ramon, Rocio de Unzurrunzaga, Betina Nishishinya, Giacomo Lucenteforte, Miguel García, David Barastegui, Itziar Unzueta, Antonio Arcalis, Ramon Cugat

## Abstract

**Introduction:** Musculoskeletal pain is a common affection due to ageing, sedentarism and injuries. The objective of this trial is to prove efficacy of a natural topical composition containing *Arnica montana, Hypericum perforatum, Calendula officinalis, Melaleuca sp*. and menthol in pain management in adults with acute or chronic pain.

**Methods:** This randomized, double-blinded and placebo-controlled trial included 200 patients with musculoskeletal pain, 100 in the intervention group receiving the topical formula and 100 in the placebo group, receiving a similar formula without active ingredients. The products were applied topically twice daily for 14 days in affected areas. Immediate pain alleviation and stiffness perception were monitored for two hours at days 0, 7 at 14. Pain reduction and recovery perception upon sustained application were assessed after 7 and 14 days.

**Results:** Intervention immediately reduced pain and stiffness at rest and in motion 30 minutes after application and kept being superior to placebo in all short-term timepoints (*p* < 0.05). Immediate pain reduction was maintained even at late stages of recovery. A two-week sustained intervention resulted in significant pain reduction and improvement in recovery perception. Even if both groups reached statistical significance with respect to baseline due to spontaneous lesion recovery, a significantly improved recovery was reported in the intervention group with respect to placebo.

**Conclusions:** Intervention was found to reduce pain and stiffness upon minutes of its application and to improve pain and mobility over the 14 days of treatment, showing benefits both for immediate alleviation and for longer term recovery.

**Level of Evidence:** Therapeutic Level I

## 1. Introduction

Well-being is understood as the absence of limitations or debilitating conditions that make impossible to enjoy a full and satisfactory life. Among these limitations, a reduced or impaired motility because of painful and inflammatory processes (acute and chronic), can greatly disturb a vital and optimistic attitude.

Chronic pain affects between one-third and one-half of the population only in the UK [1], and around 20.4% in the US [2]. It is likely to increase with population ageing and tends to be more common in women [3]. Ageing is translated in an increase in the incidence and span of chronic illnesses, including osteoarthritis, [4] fibromyalgia, [5] lower back pain, [6] non-arthritis joint pain or carpal tunnel syndrome [7], among others. In addition, muscular and joints pain unrelated to any specific disease also tends to appear with age [8]. Beyond that, current lifestyle frequently involves stressful jobs, eventful agendas and non-stop activity either in professional or leisure time. These facts extend the prevalence of chronic pain to younger segments of the population and to people not affected by previous disease.

Sedentary lifestyle and computer-based jobs represent a common cause of muscular contractures and pain, especially in the back and neck areas [9,10]. Another important contributor to chronic pain is perceived stress and anxiety response systems [11–13]. As an emergent antagonist phenomenon, increasing awareness of a healthy lifestyle has entailed a rise in the popularity of physical exercise in the last decades. As an example, the number of running events finishers in the U.S. stabilized in 2014 at almost 20 million people, with figures having been on the rise for the previous 25 years [14]. The popularization of exercise programs such as CrossFit [15] illustrate that the trend is directed not only towards a rise in practice, but also in intensity. Thus, the increase in the practice of high-intensity physical activity and the alternation between sedentary and exercise periods involve an increase in inflammatory and acute painful episodes, being tendinitis and joint lesions the major problem, followed by sprains or bruises [16].

The pharmacological management of these conditions involves nonsteroidal anti-inflammatory drugs (NSAIDs), local corticosteroids, acetaminophen, chondroitin sulfate or even opioids [7,17,18]. These are effective drugs but with serious side effects in some cases, especially in the long-term utilization [19]. Severe drawbacks related to the prolonged utilization of these drugs include drug ineffectiveness, toxicity of certain agents, hypersensitivity, gastrointestinal hemorrhage, nauseas and even fatal outcomes including death and suicide. Importantly, increased abuse of painkillers has been reported in several countries, exceeding tobacco consumption and being considered a public health problem [20–22]. Furthermore, the use of painkillers should be avoided in sensitive population groups including children and pregnant women, who are equally exposed to sustained or acute pain episodes.

Alternative solutions of greater safety but equivalent rapid relief and recovery effectiveness are a necessity. In this sense, the most useful approach are natural products, without tolerability concerns, compatible with other interventions and providing a rapid effect. These generalize access to effective treatment to both patients and physiotherapy professionals. Considering the well-established beneficial effects of physiotherapy in handling chronic pain [23], professionals need effective and safe solutions that they can use without concerns of side effects or pharmacological interactions, since they may not have access to the full medical record of the patients. In turn, the ability of handling self-treatment without concern increases the quality of life in patients suffering from pain-involving conditions.

This study evaluates the use of a topical cream composed by natural extracts with complementary and synergistic effects for pain management. *Menthol* reduces pain and increases blood flow, warming up the muscle and enhancing absorption of the rest of the extracts [24–28]. *Arnica montana* reduces pain, has anti-inflammatory effects and potentiates tissue repair [29,30]. *Hypericum perforatum* provides anti-inflammatory effect and drives tissue regeneration and scarring [31–34]. *Calendula officinalis* reduces swelling and distension, boosts healing of mild injuries and prevents infection [35–38]. Finally, *Melaleuca alternifolia* complements the anti-inflammatory effect and acts as a natural preservative due to its potent antimicrobial effect [39– 42].

The synergistic combination of these natural ingredients may provide a convenient solution for the management acute or chronic pain. The aim of this study is to assess the benefits of the topical formulation in reducing musculoskeletal pain, both as an immediate relief of pain and as a solution for injury recovery.

## 2. Methods

### 2.1. Study design

This prospective double-blinded, randomized, placebo-controlled, multi-centric clinical trial was conducted in the Rehabilitation Center of the Quironsalud Hospital of Barcelona and other centers of the Quironsalud network in Spain.

The main objective of the study was to demonstrate efficacy of the topical cream Fisiocrem^®^ to reduce perceived pain in patients suffering from moderate or severe musculoskeletal pain in the short and medium term.

Secondary outcomes assessed efficacy in reducing immediate pain, improving joint mobility and flexibility and evaluated recovery perception.

### 2.2. Subjects

Target sample size was n = 200 patients, 100 in the intervention group and 100 in the control group, allocated on a 1 to 1 ratio. Inclusion criteria were as follows: men or women above 18 years old, with acute or chronic musculoskeletal pain scoring above 4 in the Visual Analogue Scale (VAS) and with a diagnostic of either tendinopathy, vertebral algias, sprains or symptomatic osteoarthritis. Exclusion criteria were a diagnostic in either neuropathic chronic pain, fibromyalgia or neoplasia, or an allergy to any ingredient in the formula. Sample size was calculated by accepting an alpha risk of 0.05 and a beta risk of 0.2. In a two-sided test and based on previously reported results with NSAIDs, 200 subjects are necessary to find a statistically significant difference in the proportion of pain reduction between groups. No drop-out rate has been considered.

### 2.3. Materials

The study product was Fisiocrem^®^, a topical cream composed of natural ingredients for the management of pain and inflammation by massaging the affected area (i.e. muscles, joints and tendons). Patients were randomly assigned, using the Excel (Microsoft) RAND function, into 2 groups: treatment group (Fisiocrem®) and placebo group, a topical cream with similar characteristics and aspect, without active ingredients.

Allocation was concealed from the recruiter and participants.

### 2.4. Procedure

The study product was given in visit 1 (day 0), and during the next 14 days it was applied twice per day to the muscular or joint areas with moderate or severe pain, while exerting a light massage. Its effects were monitored up to two hours after application in three programmed visits (days 0, 7 and 14).

### 2.5. Study outcomes

The primary endpoint was defined as pain reduction after intervention, considering both pain at rest and in motion. This was monitored at two timescales, defined as immediate pain alleviation and sustained pain reduction. The former was monitored using a Faces Pain Scale obtained at 0.5, 1 and 2 hours after product application in each visit. The latter was assessed by subjects completing a VAS two hours after the intervention at each visit day. Both the Faces Pain Scale and the VAS evaluate pain with scores ranging from 0 to 10, with higher values indicating more pain.

Secondary endpoints were also assessed at the immediate and sustained timescales. In each visit day, the immediate intervention effects on stiffness (both at rest and in motion) were assessed at 0.5, 1 and 2 hours using a stiffness scale, being (0) no stiffness, (1) slight stiffness, (2) moderate stiffness and (3) severe stiffness.

Sustained intervention effects were monitored two hours after product application at each visit day. Joint mobility was assessed by evaluating passive joint balance in a scale ranging from 1 to 4, with the lowest score (1) corresponding to free mobility, (2) partial limitation, (3) moderate limitation and (4) severe limitation. Global perception of recovery after 7 and 14 days was measured using a Likert scale, in which (1) meant fully recovered, (2) much better, (3) better, (4) same as before and (5) worse than before.

Instructions of recording adverse effects (if any) were given to the investigators.

### 2.6. Statistical analysis

The categorical endpoints (perception of pain reduction, perception of stiffness, recovery perception and joint mobility or stiffness) were analyzed using a cumulative linear mixed model. The significance of the effect of immediate product application as well as of one and two weeks of sustained intervention on the response endpoints was evaluated through Wald tests. A significance value of 0,05 (95% confidence) was established for all measurements. The analysis of clinical efficacy has been carried out with the analysis of efficacy in ITT population, including 200 patients (n = 100 per group).

### 2.7. Ethical considerations

The study was conducted in accordance with the Declaration of Helsinki, ethical standards, current legislation and GCPs. The study was approved by the Ethics Committee of the Quironsalud Group, and informed consent was obtained from all patients prior to their enrolment. Patients were informed that they could quit the study at any time and for any reason.

#### Trial registration

This trial was approved by a local Ethical Committee with an Ethical approval sanctioned number 58/2019. This trial is registered in ClinicalTRials.gov PRS with registration number NCT04683263, as of 24^th^ December 2020 (retrospectively registered) with url https://www.clinicaltrials.gov/ct2/show/NCT04683263.

## 3. Results

### 3.1. Participants

200 patients were assessed for eligibility and randomized into the intervention (n = 100) and placebo (n = 100) groups. 12 volunteers did not attend visit 1, which resulted in 98 patients in the study group and 90 patients in the placebo group. Statistical analysis was performed on an ITT protocol, including 100 patients per group.

### 3.2. Baseline data

No differences between groups with regards to demographic data, and baseline data of pain and stiffness before the intervention were found (Table 1).

**Table 1.**
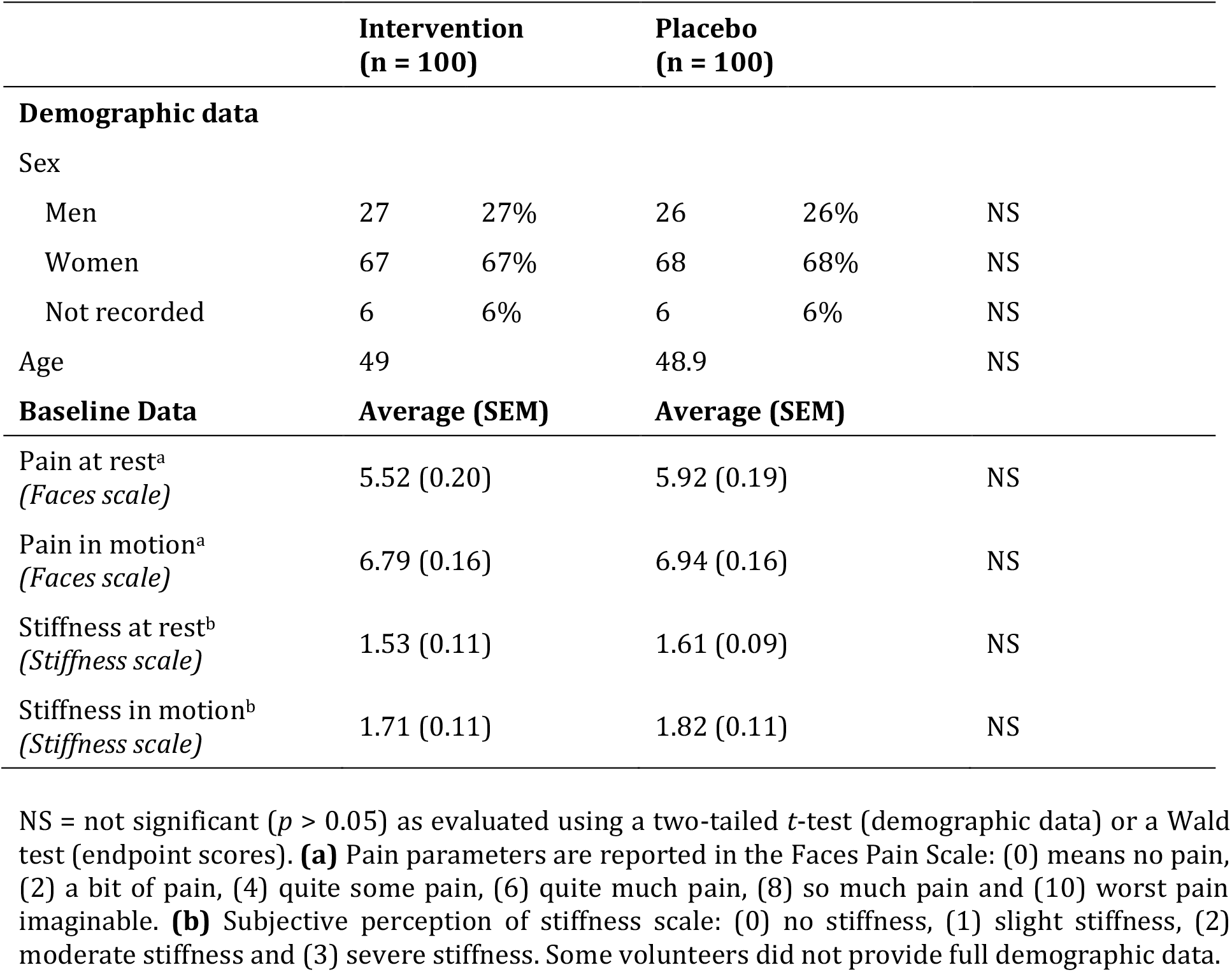
Baseline data for intervention and placebo groups:

### 3.3. Alleviation of symptoms in the short-term

#### 3.3.1. Immediate alleviation of pain at rest

Significant differences between the intervention and placebo groups were found after 30 min (25% variation), 1 hours (27%) and 2 hours (27%) on the first day of application (Figure 1A). After one week, significant differences were already present before applying the formulas (12%) due to a sustained pain reduction effect (see below). The differences between the two groups increased after 30 min (30%), 1 hour (33%) and 2 hours (34%). After two weeks, the group difference was even more obvious before product application (26%), after 30 min (43%), 1 hour (48%) and 2 hours (48%).

**Figure 1.**
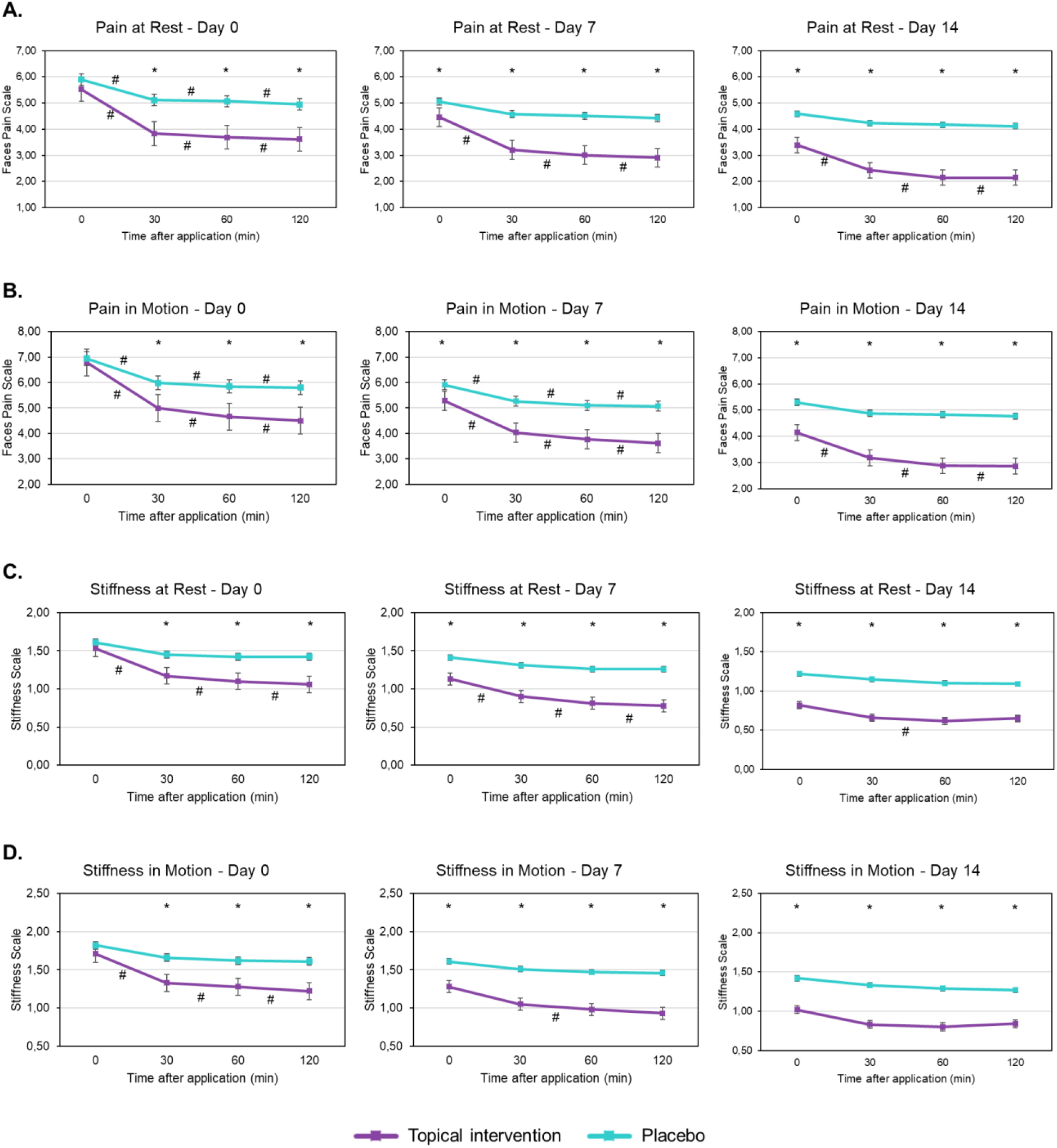
Assessment of immediate alleviation after 30 min, 1h or 2h of the application of the topical intervention or placebo. **A**. Assessment of the immediate alleviation of pain at rest and **B**. in motion using the Faces pain Scale, where (0) means no pain, (2) a bit of pain, (4) quite some pain, (6) quite much pain, (8) so much pain and (10) worst pain imaginable. **C**. Assessment of the immediate effects on stiffness perception at rest and in motion by subjective response to a stiffness scale, where (0) no stiffness, (1) slight stiffness, (2) moderate stiffness and (3) severe stiffness. \**p* < 0.05 between intervention and placebo. # *p* < 0.05 between timepoints. Error bars indicate the SEM.

Within each group, differences among the 0, 0.5, 1 and 2 hours timepoints were always significant in the intervention group, while no statistical significance was observed for the placebo group at days 7 and 14. This indicates sustained effectivity of the formula even at advanced stages of lesion recovery.

#### 3.3.2. Immediate alleviation of pain in motion

Variations between the intervention and placebo groups were already significant after 30 min (17%), 1 hours (20%) and 2 hours (22%) on the first day of intervention (Figure 1B). After one and two weeks, a sustained pain reduction effect (see below) implied that significant differences were already present before applying the formula (11% and 22% for days 7 and 14, respectively). Such differences were increased after 30 min (23% and 35%) 1 hour (26% and 41%) and 2 hours (19% and 40%).

Within each group, while differences among the monitored timepoints were significant at all visits for the intervention group, at day 14 they were not for the placebo group. Again, this supports the sustained effectivity of the formula for pain alleviation.

#### 3.3.3. Immediate effects on stiffness at rest

On the first visit, significant differences between the intervention and the placebo groups were already found 30 min after product application (19% of variation), and increased after 1 hour (22%) and 2 hours (25%) (Figure 1C). At days 7 and 14, significant differences were present before application (20% and 33%), thus indicating sustained stiffness reduction effectivity of the formula. Differences increased after 30 min (31% and 42% for days 7 and 14, respectively) 1 hour (36% and 44%) and 2 hours (38% and 40%).

Of note, differences among timepoints were not significant for the placebo group at any study stage. Conversely, a significant stiffness reduction with time was observed in the intervention group at days 0 and 7. At day 14, significant differences were found only after 1 hour of product application, probably due to the reduced improvement margin left for lesions in an advanced recovery stage.

#### 3.3.4. Immediate effects on stiffness in motion

On day 0, significant differences between groups were observed already after 30 min (20%), which increased after 1 hour (21%) and 2 hours (24%). After one and two weeks of treatment, significant differences were already observed at time 0 (20% and 28% for days 7 and 14, respectively) due to the sustained effectivity of the formula at reducing the perception of stiffness in motion. Differences between groups increased after 30 min (31% and 38%), 1 hour (34% and 37%) and 2 hours (36% and 34%).

Within groups, timepoint differences were never significant for the placebo group, while in the intervention group they were consistently significant throughout day 0 and after 1 hour of product application on day 7. Again, this indicates that the formula is effective at reducing stiffness on early lesion recovery stages (Figure 1D).

### 3.4. Effects of sustained intervention

The effects of the sustained intervention (twice a day for 14 days) are summarized in Table 2.

**Table 2.**
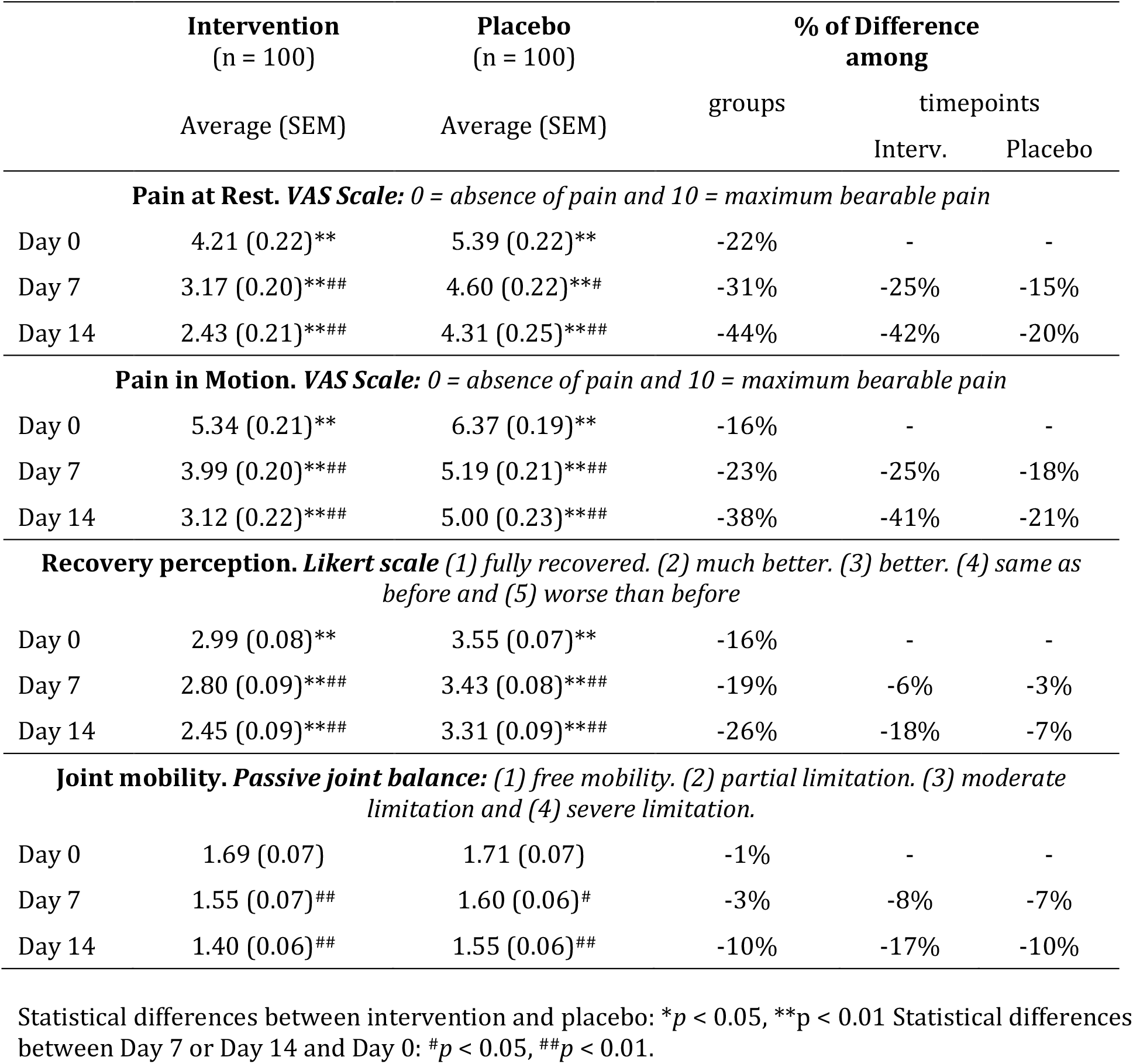
Differences in pain, mobility, and recovery perception among groups (intervention vs. placebo) and among timepoints (Day 7 and Day 14 vs. Day 0).

Pain (both at rest and in motion) was evaluated using a VAS scale two hours after intervention at each visit day. For both pain variables, differences between the intervention and the placebo groups were already present two hours after the first application (day 0) and kept being consistently significant throughout the time period of the study, thus evidencing the superior effect of the formula. Within each group, significant differences between the day 0, 7 and 14 timepoints reflect the progression of lesion recovery.

Joint mobility was also evaluated after two hours of intervention at each visit day, using a passive joint balance scale. Timepoint differences within groups reflected lesion recovery progression, even if differences between groups were not significant.

Finally, a Likert scale was used to measure the recovery perception. Differences between groups were consistently significant throughout the study, indicating formula superiority.

### 3.5. Safety and tolerability

No adverse effects were observed throughout the study. Volunteers reported an overall good perception of the treatment and no compliance or tolerability issues were detected.

## 4. Discussion

The design and characteristics of the study respond to the interest of demonstrating the analgesic and reparative functionalities of Fisiocrem^®^, formulated with natural plant extracts. According to the intrinsic product characteristics, its application route and the final objective - accelerate muscle and articular lesions recovery, which takes place naturally over time - it is essential to use a placebo group with which to neutralize possible biases attributable to psychological and physiological factors.

In accordance with the contrasted properties of its active ingredients, Fisiocrem^®^ is formulated to favour lesion recovery. It provides a pleasant sensation of relief and well-being immediately after its application, thus meeting an heterogenous need in a wide range of patients. For this reason, inclusion criteria were designed to enrol patients suffering ailments in multiple anatomical locations, chronically or in acute timeframe.

The primary objective of the study was to assess the ability of Fisiocrem^®^ to reduce pain during lesion recovery or improve debilitating chronic disfunctions. The product has been formulated to reduce inflammation and improve tissue repair through the synergic action of its natural components (*Menthol, Amica Montana, Hypericum perforatum, Calendula officinali* and *Melaleuca alternifoliaii*).

This study has been designed to monitor the evolution of lesions and injuries at two timescales while avoiding or minimizing masking effects. On the short timescale, our results show the immediate efficacy of Fisiocrem^®^ in alleviating both pain at rest and in movement (Figure 1A and B). On the long term, a sustained application of Fisiocrem^®^ for two weeks results in a superior alleviation of pain when compared to placebo (Table 2). Together with significant improvements in stiffness alleviation in the short term (Figure 1C and D) and recovery perception in the long term (Table 2), these data provide a clear vision of the statistical but also physiotherapeutic relevance of the results in all the parameters tested. Even when the placebo group is experiencing a good percentage of improvement due to spontaneous recovery, in all cases Fisiocrem® exceeds the placebo results becoming a superior solution.

To fully understand the range of applications of Fisiocrem^®^, the fast pain relief becomes essential. This property presents a double value. On the one hand, to encourage patient adherence to treatment and thus facilitate its reparative action and recovery from injury. On the other hand, this provides patients and physiotherapy professionals with a solution to improve highly disabling pain situations, constituting an alternative to pharmacological principles but with a much safer profile. Thus, the safety profile of Fisiocrem^®^, with no limit in the number of daily applications, offers a *quantum satis* solution for pain relief. On this regard, as summarized in Figure 1, the simple application of the placebo product by means of a massage improves the subjective perception of the patients in each one of the analysed parameters from 30 to 120 minutes, which can be explained considering natural lesion recovery. However, the efficacy for rapid relief of the extracts formulated in Fisiocrem® (menthol and *Arnica montana*) presents better results than the placebo in all days and times tested. This observation reinforces the product proposal as an alternative to systemic, safety-limited, pharmacologic treatments such as NSAIDs. Concomitantly, the immediate reduction of pain or stiffness perception after product application is significant regardless of spontaneous lesion recovery over time only in the intervention group, since improvement reported by the patients receiving the treatment keeps its clinical relevance along the trial. This highlights the efficacy of Fisiocrem^®^ at reducing pain even at advanced stages of lesion recovery.

Along with immediate symptom relief, our results show that Fisiocrem^®^ efficiently reduces pain and improves recovery perception when applied in a sustained manner for two weeks. Thus, this provides patients with a solution to self-treat injuries on a daily basis, while it serves physiotherapy professionals as a reliable tool to be applied alongside professional massage in discrete visits during treatment.

While the present study provides evidence of the benefits of Fisiocrem^®^ for pain management, some aspects of the study design may limit its interpretation. Regarding the studied cohorts, despite being integrated by patients with heterogeneous characteristics the study groups are comparable according to baseline data. However, a classification of subjects into groups of acute or chronic pain would allow an independent analysis of the recovery and short-term perception in each group of patients, which could help identify benefits and particularities of the product in chronic or acute pain or lesions, respectively. Future trials enrolling patients affected by chronic musculoskeletal pain or debilitating conditions requiring larger periods of recuperation should include longer periods of monitoring, allowing to determine and identify the limit of recovery attributable to the product of application and not to the spontaneous resolution of the injury. Some limitations also arise with regards to the interpretation of the fast recovery after product application. In line with the previous observation, a longer follow-up than 2 hours after the application of Fisiocrem^®^ would help determine the duration of its relief effect and would make possible to even consider a head-to-head comparison with pharmacologic treatments.

Fisiocrem^®^ has been shown to be safe and efficacious for pain management and recovery of lesions of different origin. Its effects over time make it an ideal natural approach for the improvement of pain and disability that can be safely applied home by the patient on a daily basis.

## 5. Conclusions

The immediate alleviation of pain and stiffness conferred by Fisiocrem^®^ implicates that the product is an optimal solution both for reducing the pain caused by the lesion itself but also for alleviating the discomfort that the therapy may cause. In addition, and due to its dual action, alleviation and recovery, Fisiocrem® meets the requirements for home, professional and combined use.

## Data Availability

All data produced in the present work are contained in the manuscript

## Abbreviations

NSAIDS: non-steroidal anti-inflammatory drugs
SEM: standard error of the mean
VAS: Visual Analog Scale

## Acknowledgements

Authors dedicate this work to Dr. Silvia Ramón (R.I.P.), who directed the study as Principal Investigator and contributed to the preparation and revision of the manuscript. Authors thank Dr. Jordi Cuñé and Dr. Maria Tintoré for manuscript writing and results analysis assistance.

